# Feasibility and visualization of deep learning detection and classification of inferior vena cava filters

**DOI:** 10.1101/2020.06.06.20124321

**Authors:** Brian J. Park, Vlasios S. Sotirchos, Jason Adleberg, S. William Stavropoulos, Tessa S. Cook, Stephen J. Hunt

## Abstract

**Purpose:** This study assesses the feasibility of deep learning detection and classification of 3 retrievable inferior vena cava filters with similar radiographic appearances and emphasizes the importance of visualization methods to confirm proper detection and classification.

**Materials and Methods:** The fast.ai library with ResNet-34 architecture was used to train a deep learning classification model. A total of 442 fluoroscopic images (N=144 patients) from inferior vena cava filter placement or removal were collected. Following image preprocessing, the training set included 382 images (110 Celect, 149 Denali, 123 Günther Tulip), of which 80% were used for training and 20% for validation. Data augmentation was performed for regularization. A random test set of 60 images (20 images of each filter type), not included in the training or validation set, was used for evaluation. Total accuracy and receiver operating characteristic area under the curve were used to evaluate performance. Feature heatmaps were visualized using guided backpropagation and gradient-weighted class activation mapping.

**Results:** The overall accuracy was 80.2% with mean receiver operating characteristic area under the curve of 0.96 for the validation set (N=76), and 85.0% with mean receiver operating characteristic area under the curve of 0.94 for the test set (N=60). Two visualization methods were used to assess correct filter detection and classification.

**Conclusions:** A deep learning model can be used to automatically detect and accurately classify inferior vena cava filters on radiographic images. Visualization techniques should be utilized to ensure deep learning models function as intended.

## Background

Over the past few years, advances in machine learning (ML) have led to efforts in developing clinical applications in medicine and especially in radiology. These applications have started to be utilized in radiology departments for workflow optimization, pathology identification, and study triaging [1, 2]. Although interventional radiology practices have not been significantly impacted to date by these developments, potential applications have been identified, such as diagnosis assistance, patient selection for procedures, clinical decision support, and treatment response classification [3, 4].

One task that could feasibly be facilitated by ML algorithms is the identification of inferior vena cava (IVC) filters on radiographic images, owing to their fixed filter design and metallic density. With the increased awareness of IVC filter complications, knowledge of the exact filter type is important for providers, diagnostic radiologists, and interventional radiologists in determining appropriate patient management [5-7]. Plain radiography is often the first step in evaluating patients who have a history of filter placement. The increased spatial resolution on radiographic images compared to CT can assist in determining filter type when placement records are not available, as well as allow for the detection of fractured filter components [8].

Many IVC filters have similar designs. The resources and costs for training and educating diagnostic radiologists, as well as the extra image interpretation and dictation time needed to classify each filter on a radiographic image, may not be a cost-effective strategy. A deep learning convolutional neural network (CNN) has been recently demonstrated to successfully classify 14 different types of filters with high sensitivity and specificity using cropped, high-quality radiographic images [9]. However, in clinical practice, radiographs are not routinely manipulated to focus specifically on the filter, and often these images are suboptimal due to patient-related factors and the presence of overlying radiopaque objects. Furthermore, filters may be distorted or have fractured components, adding complexity to the task of pattern recognition.

In this study, the feasibility of deep learning for both detection and classification is demonstrated using radiographic images of three distinct but similarly-appearing retrievable IVC filters: Celect (Cook Medical, Bloomington, IN), Denali (Bard Medical, Covington, GA), and Günther Tulip (Cook Medical, Bloomington, IN). Visualization of relevant regions and features used for classification by the deep learning model was performed using two different visualization techniques, emphasizing the complexity of deep learning CNNs and challenges in troubleshooting proper machine functioning [10].

## Methods

This study was reviewed and approved by the Institutional Review Board. A deep learning classification algorithm based on supervised learning using ResNet-34 architecture was trained using the fast.ai library (v1.0.54) on a workstation equipped with Core i7-8700K, 32GB RAM, and NVIDIA Titan XP GPU. Fast.ai is a free and popular deep learning library and application programming interface that is widely used in academia and industry for its ease of use and state-of-the-art results [11]. It is noteworthy that most deep learning image classification networks are pre-trained on colored RGB images [12]. When applied to radiological images that are greyscale, all 3 color channels (red, green, and blue) are still utilized. Radiographic images from IVC filter placement or removal from 144 patients during February 2017-December 2018 were exported from PACS, anonymized, and converted to grayscale jpg format. These images included digital spot radiographs, stored fluoroscopic images, and images from rotational cavography. The type of filter was extracted from the procedural report. Images were cropped to remove text labels and edge collimation only; but more specifically, images were not cropped to center around the filter. Following preprocessing, the total number of images was 442, varying from entire views of the abdomen to fluoroscopically magnified views of the filter (Fig. 1).

**Figure 1.**
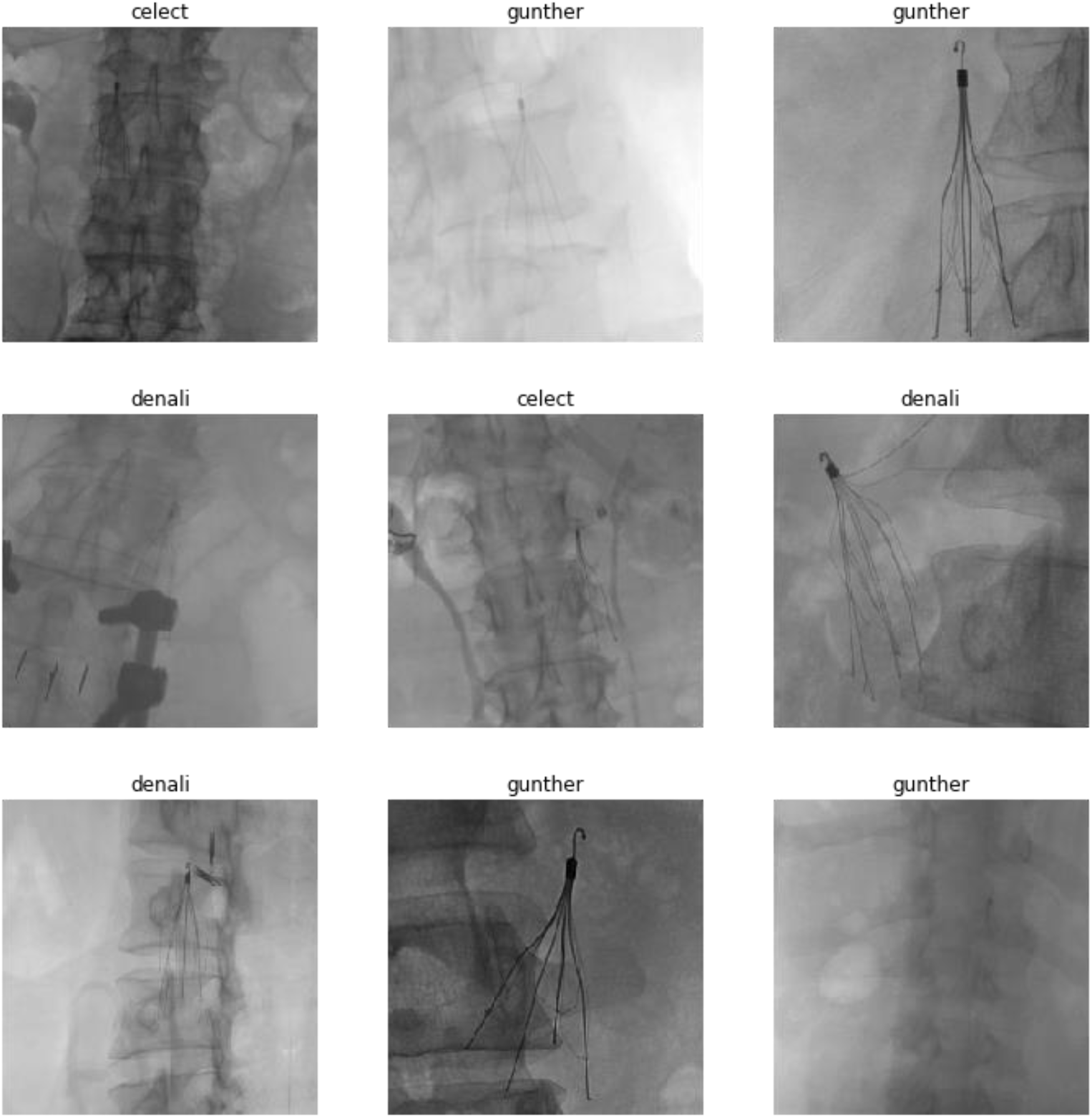
Sample set of images within the dataset. Images were not selectively cropped to focus on the filter. Variable image quality based on image exposure, patient positioning, and body habitus; filter distortions; and overlying radiodense objects are present.

The training set included 382 images (110 Celect, 149 Denali, 123 Günther Tulip), of which 80% were used for training and 20% for validation. Data augmentation including horizontal flip, random rotation, windowing, zooming, and warping was performed for regularization using default fast.ai library settings. A random test set of 60 images (20 images of each filter), not included in the training or validation set, was used for evaluation. Total accuracy and receiver operating characteristic area under the curve (ROC AUC) were used to evaluate performance. Overall macro-average of the ROC AUCs was calculated. Feature heatmaps were visualized using guided backpropagation (GBP) [13] and gradient-weighted class activation mapping (Grad-CAM) [14]. Grad-CAM provides a coarse heatmap of regions on the image that had the greatest influence for classification. GBP provides an overview of all the features detected by the network, such as edges, textures, curves, and shapes, with colors representing the relative strength of each weight from the red, green, and blue channels.

## Results

The model was trained in 20 epochs with a total compute time of 21 seconds. Following training, the total accuracy on the validation set was 80.2% (15/22 Celect, 20/25 Denali, 26/29 Günther Tulip) with mean ROC AUC of 0.96 (Fig. 2). Evaluation on the test set yielded total accuracy of 85.0% (Table 1) with mean ROC AUC of 0.94 (Fig. 2). Data visualization demonstrated detection and misclassification errors, including correct filter classification despite failing to detect the filter on the image (Figs. 3-6).

**Table 1.**
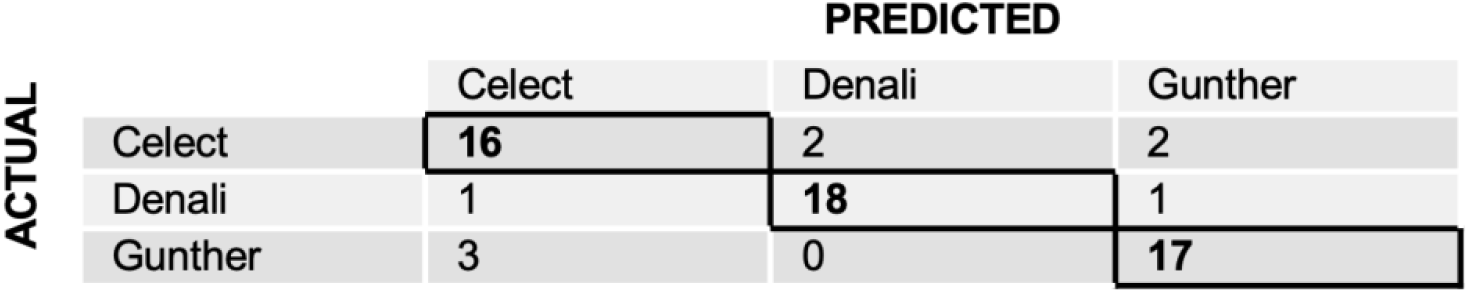
Confusion Matrix for Test Set (N=60)

**Figure 2.**
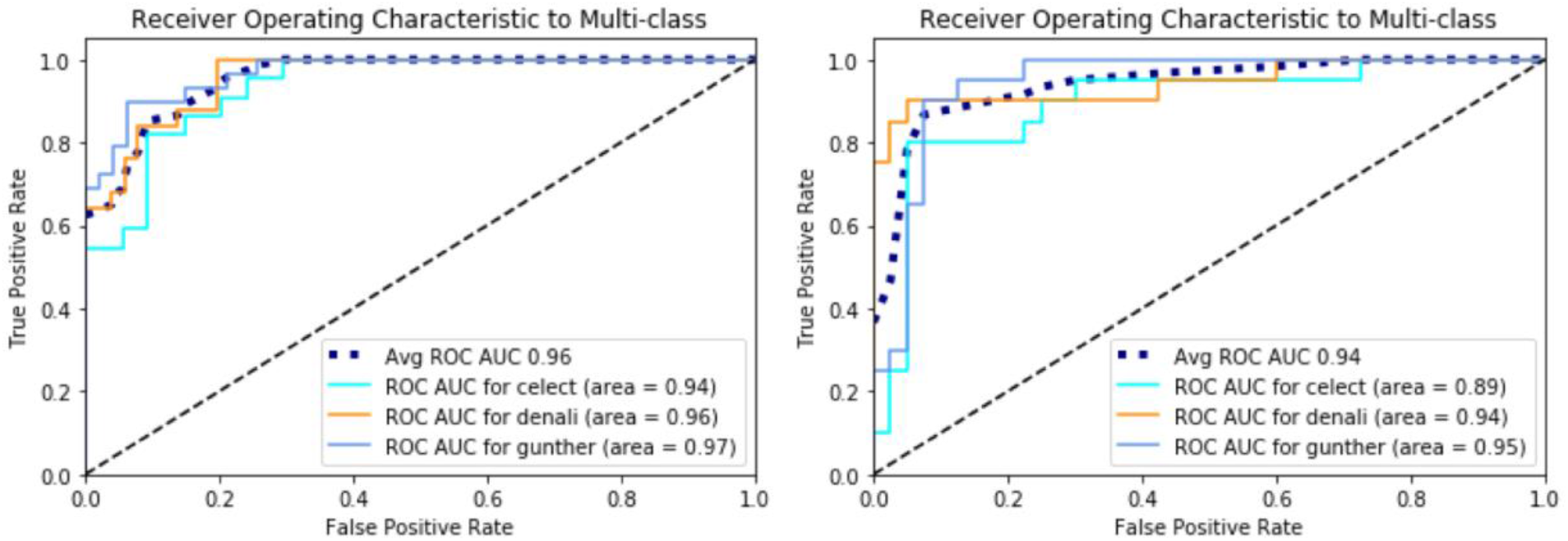
*Left*: ROC AUC of each filter in validation set (N=76) with mean of 0.96. *Right*: ROC AUC of each filter in test set (N=60) with mean of 0.94.

**Figure 3.**
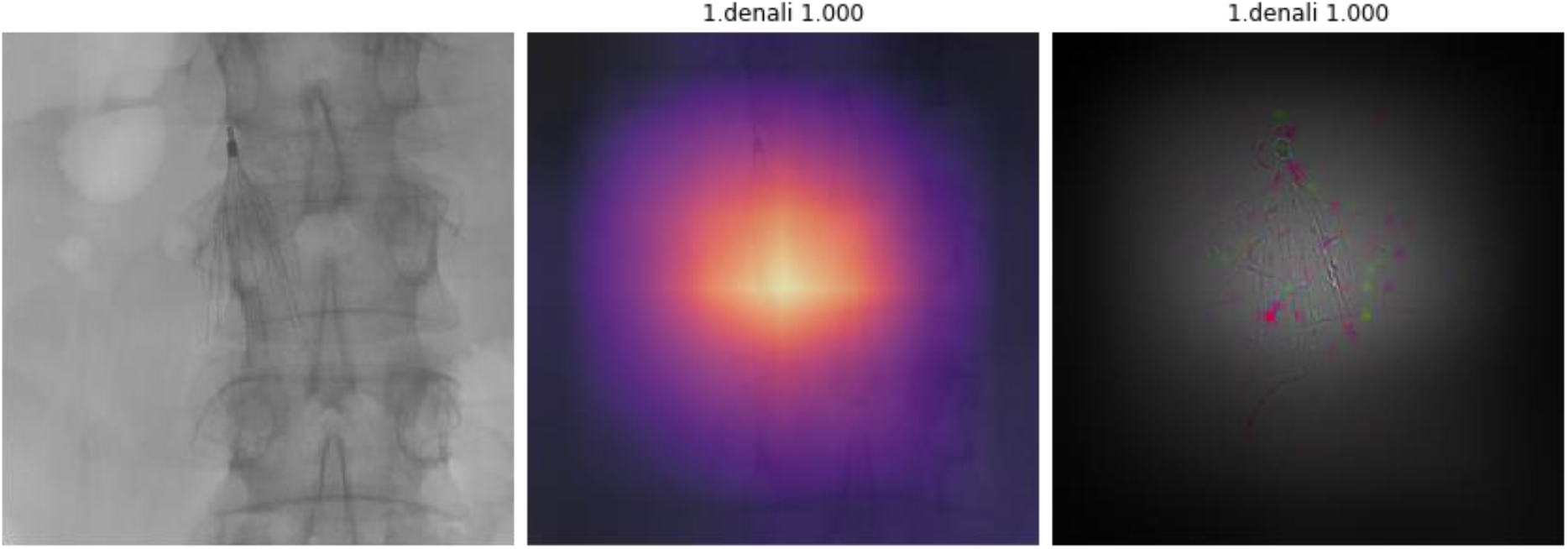
Correct detection and identification (predicted Denali, actual Denali). Majority in the test set were in this category. Left is raw image, middle is Grad-CAM, and right is GBP. Grad-CAM and GBP both show correct localization of the filter and detection of filter features. The machine gave a 100% probability that the filter was a Denali.

## Discussion

The results show the feasibility of deep learning CNNs to accurately classify various types of IVC filters with average ROC AUCs of 0.94-0.96. Model training with the fast.ai library was very fast, taking only 21 seconds to train 382 images. Feature visualization for the majority of test cases confirmed correct filter detection and classification, as shown in Fig. 3. Interestingly, visualization also indicated that a disproportionate number of Celect filter images had the presence of EKG leads, which were used to classify Celect filters from Denali or Günther Tulip. In Fig. 4, the model missed detection of the filter but correctly classified the Celect filter with 93.4% probability through features from the EKG leads. Correct filter detection but misclassification, as shown in Fig. 5, is likely due to the limited number of features detected or small sample size. Fig. 6 reveals the importance of using more than one visualization technique. Although the machine missed detection and classification of the filter, visualization shows that some features of the filter were in fact activated, such as the tip.

**Figure 4.**
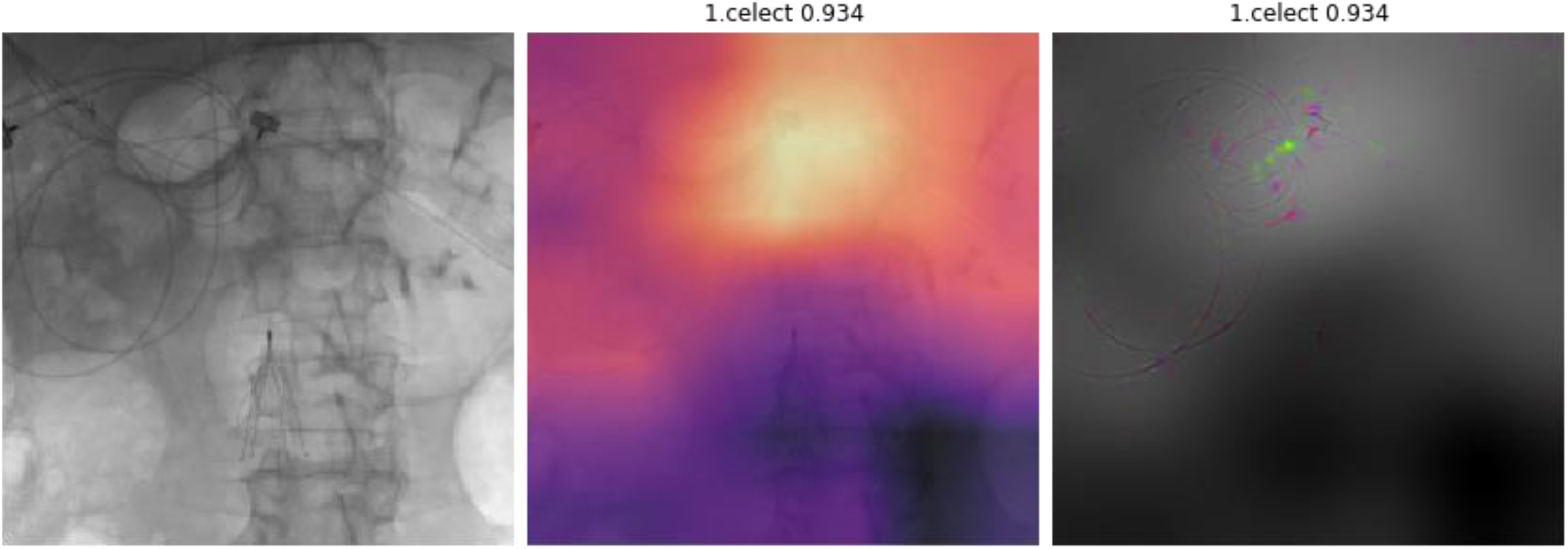
Missed detection but correct classification (predicted Celect, actual Celect). Left is raw image, middle is Grad-CAM, and right is GBP. Grad-CAM and GBP both show incorrect localization of the filter and detection of features related to the ancillary EKG leads. The machine gave a 93.4% probability that the filter was a Celect.

**Figure 5.**
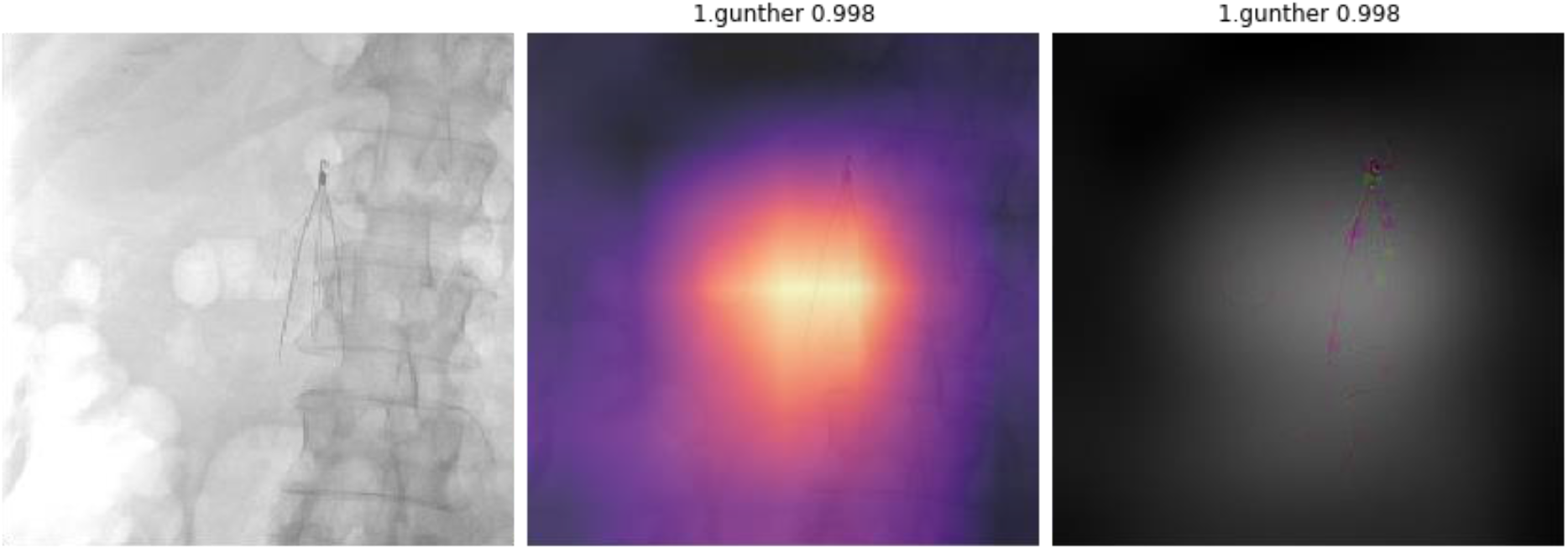
Correct detection but misclassification (predicted Günther Tulip, actual Denali). Left is raw image, middle is Grad-CAM, and right is GBP. Grad-CAM and GBP both show correct localization of the filter and detection of filter features, but the machine gave a 99.8% probability that the filter was a Günther Tulip.

**Figure 6.**
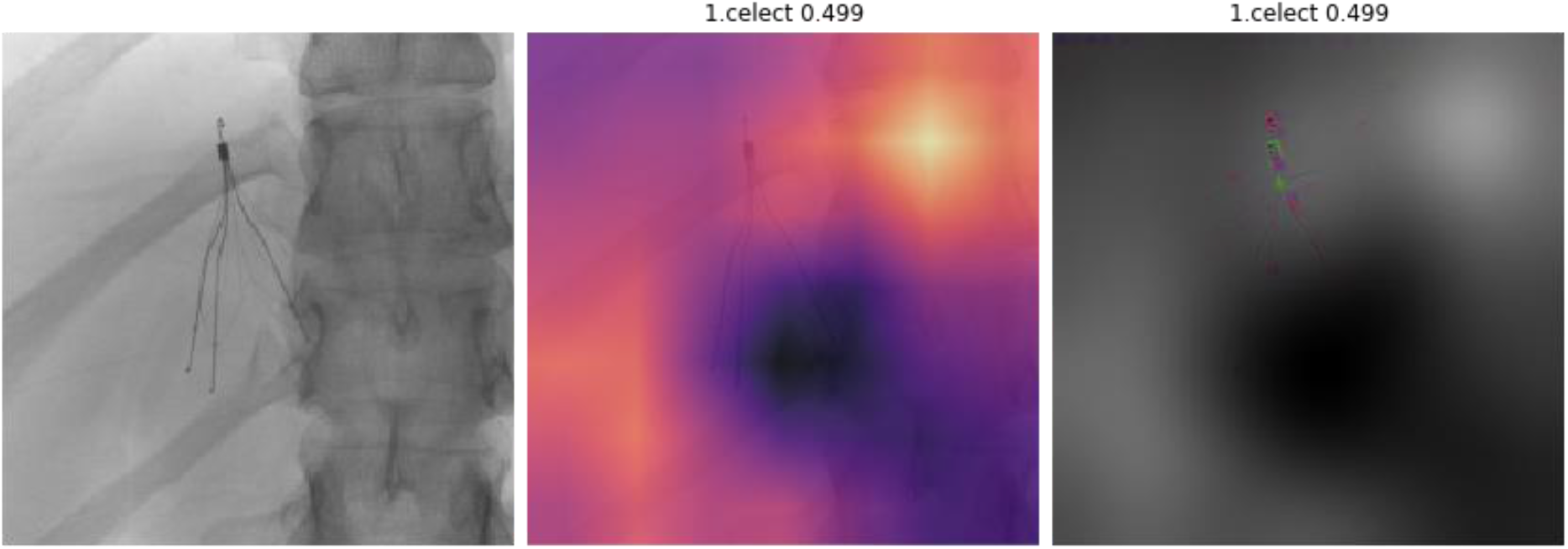
Missed detection and misclassification (predicted Celect, actual Günther Tulip). Left is raw image, middle is Grad-CAM, and right is GBP. Grad-CAM primarily shows localization on a vertebral body, but GBP shows some detection of filter features like the tip. The machine gave a 49.9% probability that the filter was a Celect.

Several methods exist to visualize features activated, or emphasized, by a deep learning network to ensure proper functioning. Two common visualization techniques were implemented, both of which are gradient-based methods: Grad-CAM [14] and GBP [13]. Grad-CAM examines gradient information flowing forward into the final convolutional layer of a neural network, and generates a localized, coarse heatmap of regions in the image that had the largest influence in the model’s ability to discriminate between classes. Thus, Grad-CAM can be used as a coarse indicator to see if the machine identifies the location of the filter on the image. On the other hand, GBP highlights more granular, fine-grained features that were used to classify the image. Rather than focusing primarily on information flowing into the final convolutional layer, gradient information is propagated backwards through all the layers of the neural network in order to discern all contributing features from the original image that were used to make a decision. In essence, although visualization techniques provide insights into the inner workings of a deep learning classification algorithm, these techniques are not able to explain why certain cases fail, like in Fig. 6, with missed detection and missed classification but with some features of the filter activated within the network. The complexity of deep learning CNNs, with millions of parameters and activations, underscores the challenges of troubleshooting and debugging the network.

Ni *et al* previously demonstrated that a deep learning model can accurately classify 14 different types of filters with an overall accuracy of 97.1% (135/139) in their test set [9]. The overall accuracy was 85.0% (51/60) for the test set in this study. However, in contrast to their dataset, where images were selectively screened for good quality and minimally deformed filters, images in this study were not centered, magnified, or cropped *post hoc* to focus solely on the filter. Additionally, all image qualities and filter conditions were enrolled in this study, including distorted filters, fractured filters, and filters partially obscured by overlying radiodense objects, such as surgical clips, EKG leads, and spinal fusion hardware. For Celect filters, the machine used the presence of EKG leads for filter classification. A potential solution for this issue can be implementing additional input information via bounding boxes around the filters for model training to limit biasing from extraneous information on the images [15].

Radiographic images of the abdomen without filters were not included in this study. The absence or presence of an IVC filter is better suited for a separate machine learning algorithm altogether, either supervised or even unsupervised, as the classification question changes from one conditional statement, “What filter is this?” to 2 nested conditional statements, “Is there a filter? If so, what filter is this?” Subsequently, if a filter is present, the image can then be fed into and analyzed by a more specific filter classification algorithm. Inserting images without filters into the training set for this model would have likely diminished overall sensitivity and specificity since the finite allocation of weights for differentiating between similarly-appearing filter types would have been somewhat diluted with weights dedicated towards determining the absence or presence of a filter instead.

A major limiting factor of this study is that only 3 types of IVC filters were used. Nevertheless, this is a feasibility study demonstrating that deep learning has the potential to automatically detect and accurately classify similarly-appearing IVC filters. Additionally, visualization techniques are essential to confirm proper functioning of ML algorithms, and various techniques exist to help elucidate the workings of deep CNNs. Future plans involve the addition of other filter types as well as significantly increasing the sample size to improve accuracy and generalizability. Another area of interest is extending automated IVC filter detection and classification to diagnostic radiographic images and CT scans.

## Conclusions

Deep learning models can be used to automatically detect and accurately classify IVC filters on radiographic images, which may assist radiologists and ordering providers in filter management. Visualization techniques are useful to ensure deep learning models function as intended, but the complex interconnectivity and vastness of CNNs preclude detailed assessment and troubleshooting of situations that fail. Although this represents a feasibility study with a limited dataset, further development and a larger dataset will improve accuracy and generalizability for clinical translation and application.

## Data Availability

Data may be available upon request. No external datasets were used.

## Abbreviations

CNN: convolutional neural network
EKG: electrocardiogram
Grad-CAM: gradient-weighted class activation mapping
GBP: guided backpropagation
IVC: inferior vena cava
ML: machine learning
ROC AUC: receiver operating characteristic area under the curve

## Notes

### Competing Interest Statement

The authors have declared no competing interest.

### Funding Statement

The Titan Xp used for this research was donated by NVIDIA Corporation.

### Author Declarations

University of Pennsylvania

